# The benefits and harms of adjuvant chemotherapy for non-small cell lung cancer in patients with major comorbidities: A simulation study

**DOI:** 10.1101/2022.01.31.22270197

**Authors:** Amanda Leiter, Chung Yin Kong, Michael K. Gould, Minal S. Kale, Rajwanth R. Veluswamy, Cardinale B. Smith, Grace Mhango, Brian Z. Huang, Juan P. Wisnivesky, Keith Sigel

## Abstract

**Background:** Randomized controlled trials (RCTs) have demonstrated a survival benefit for adjuvant platinum-based chemotherapy after resection of stage IB-IIIA non-small cell lung cancer (NSCLC). The relative benefits and harms and optimal approach to treatment for NSCLC patients who have major comorbidities (chronic obstructive pulmonary disease [COPD], coronary artery disease [CAD], and congestive heart failure [CHF]) are unclear, however.

**Methods:** We used a simulation model to run in-silico comparative trials of adjuvant chemotherapy versus observation in stage IB-IIIA NSCLC in patients with comorbidities. The model estimated quality-adjusted life years (QALYs) gained by each treatment strategy stratified by age, comorbidity, and stage. The model was parameterized using outcomes and quality-of-life data from RCTs and primary analyses from large cancer databases.

**Results:** Adjuvant chemotherapy was associated with clinically significant QALY gains for all patient age/stage combinations with COPD except for patients >80 years old with stage IB cancers. For patients with CHF and stage IB disease, adjuvant chemotherapy was not advantageous; in contrast, it was associated with QALY gains for stages II-IIIA for younger patients with CHF. In general, patients with multiple comorbidities benefited less from adjuvant chemotherapy than those with single comorbidities and women with comorbidities in older age categories benefited more from adjuvant chemotherapy than their male counterparts.

**Conclusions:** Older, multimorbid patients may derive QALY gains from adjuvant chemotherapy after NSCLC surgery. These results help extend existing clinical trial data to specific unstudied, high-risk populations and may reduce the uncertainty regarding adjuvant chemotherapy use in these patients.

## Background

Lung cancer is the leading cause of cancer death in the United States, with non-small cell lung cancer (NSCLC) representing >85% of lung cancer cases (1, 2). Locoregional disease makes up approximately 35% of lung cancer cases and this proportion is projected to grow with the increased uptake of low-dose computed tomography screening (3, 4). As meaningful long-term survival is possible for patients with locoregional disease, optimizing treatment in this patient population is critical (5).

A substantial proportion of patients with locoregional NSCLC have serious comorbidities that are related to smoking. The comorbidities most commonly observed in patients with NSCLC include cardiovascular disease (20-25%) and chronic obstructive pulmonary disease (COPD; 25-50%) (6-10). Patients with NSCLC and comorbidities are less likely to receive cancer treatment concordant with medical guidelines, are more likely to experience treatment-related adverse events and have worse survival (6, 10).

Surgery is the standard treatment for locoregional NSCLC, but a large proportion of patients remain at risk of cancer recurrence after resection (11, 12). Several randomized control trials (RCTs) demonstrated a survival benefit for adjuvant platinum-based chemotherapy after resection of stage IB-IIIA NSCLC (4-5% absolute increase in 5-year survival), leading to strong recommendations for this practice in current treatment guidelines (11, 13, 14). However, patients with serious comorbidities are commonly excluded from cancer RCTs and the benefits and harms of adjuvant chemotherapy are unclear in this patient population (15-17).

The risk/benefit ratio of adjuvant chemotherapy may substantially differ in patients with comorbidities due to differences in adverse events from cancer treatment, competing mortality risks (non-lung cancer deaths), and poorer quality of life (18-22). Patients with comorbidities have higher mortality from non-lung cancer causes and as competing risks increase, the potential early adverse events associated with chemotherapy must be weighed against the long-term survival benefits of chemotherapy, which are not experienced for many years after treatment. Additionally, patients with comorbidities may have poorer baseline quality of life which might attenuate absolute improvements in quality-adjusted life years from more aggressive lung cancer treatment compared to patients without comorbidities. Understanding the potential benefits and harms of adjuvant chemotherapy in patients with comorbidities is essential for optimizing treatment outcomes and aiding clinical decision-making. In this study, we aim to determine the indications for adjuvant chemotherapy in patients with NSCLC and smoking-related comorbidities (COPD and cardiovascular diseases) by developing a simulation model to run in-silico comparative trials of platinum-based adjuvant chemotherapy versus observation in stage IB-IIIA NSCLC.

## Materials and Methods

### Simulation Model Overview

We comprehensively reviewed the literature and conducted primary analyses of large cancer datasets to assess factors unique to patients with NSCLC and comorbidities. We then incorporated this information into a novel adjuvant chemotherapy-focused treatment extension of the Lung Cancer Policy Model, a validated, comprehensive micro-simulation model of lung cancer detection, development, progression, treatment, and survival (23). This treatment-focused extension of the LCPM framework, which we will refer to as Comorbidity Lung Treatment Model (COLT-M), includes new data related to comorbidities, treatment-related adverse events, long-term survival, and NSCLC characteristics across multiple nationally-representative clinical datasets.

In COLT-M, patients >65 years old with stage IB-IIIA NSCLC and comorbidities populate the model and transition through different health states. Monthly transition probabilities were ascertained from primary analyses of large clinical datasets, clinical trial results, and a review of the literature (Table 1). After undergoing lobectomy, simulated patients were at risk for NSCLC recurrence conditional on stage and then were “randomized” to either observation or platinum-based adjuvant chemotherapy. As cisplatin-or carboplatin-based treatment regimens have similar safety and efficacy outcomes and are used in similar practice settings, the simulated adjuvant platinum-based treatment arm did not make a distinction between these types of treatments (24). Simulated patients in the adjuvant treatment arm were at risk of experiencing treatment-related complications based on age, sex, and comorbidity profiles. All simulated patients underwent monthly transition cycles until death from lung cancer or competing causes, with each cycle leading to one accumulated month of survival, which was assigned a quality-of-life weight based on treatment type, treatment complications, and comorbidities.

**Table 1.** Key Input Parameters for Developing a Microsimulation Model of Patients with Non-small Cell Lung Cancer

| Model Parameter | Definition | Value |  | Sources |
| --- | --- | --- | --- | --- |
| Response to Lung Cancer Treatment | Hazard ratios for lung cancer specific survival for adjuvant chemotherapy vs. observation | Adjuvant chemotherapy vs. observation | HR: 0.83, 95% confidence interval: 0.76-0.90 | (14) |
| Complications from Lung Cancer Treatment | Probability of complications from lung cancer treatment | Multiple complications | See Table 3 | Primary data; Table 3 |
| Mortality from Non-Lung Cancer Causes | Death rates for non-lung cancer causes by age and comorbidity | Comorbidity vs. no comorbidity | See Table 2 | Primary data; Table 2 |
|  |  | **Comorbidity** | **Utility** |  |
| Quality of Life for Patients with Comorbidities | Expected quality of life by comorbidity | CAD | -0.018 | Primary data |
|  |  | COPD | -0.021 |  |
|  |  | CHF | -0.069 |  |
| Quality of Life for Patients with Complications from Lung Cancer Treatment | Disutilities for major lung cancer treatment complications | Major complications of treatment | -0.35** | (25, 26) |
| **Quality of life returns to baseline within six months, modeled as a linear recovery. HR: hazard ratio, CAD: coronary artery disease, COPD: chronic obstructive pulmonary disease, CHF: congestive heart failure. |  |  |  |  |

This study was approved by the Icahn School of Medicine at Mount Sinai Institutional Review Board (IRB 16-00576) with a waiver of consent in compliance with the Code for Federal Regulations Title 45 Part 46.116.

### Input Data and Parameters

COLT-M parameters include mortality (both from NSCLC and competing risks), complications from treatment, and quality of life utilities. Parameters and data sources are listed in Table 1.

### Efficacy of adjuvant platinum-based chemotherapy

To estimate the benefits of adjuvant platinum-based chemotherapy on NSCLC-specific mortality, we used the results of the Lung Adjuvant Cisplatin Evaluation pooled analysis.(14) The monthly probability of NSCLC mortality was modeled based on the summary hazard ratio (HR) reported in this analysis (0.83, 95% confidence interval [CI]: 0.76-0.90). As no interactions were observed between the effects of adjuvant platinum-based therapy and age, histologic sub-type, or other patient factors in the analysis, the same HR was applied to all simulated patients.

### NSCLC-Specific and Non-NSCLC Mortality

We used population-based data from Surveillance, Epidemiology, and End-Results database linked to Medicare (SEER-Medicare) to estimate mortality from both NSCLC and competing risks (i.e., non-NSCLC causes). SEER-Medicare links SEER registry data with Medicare claims records and has information on NSCLC characteristics and outcomes, as well as allows to assess for comorbidities and complications. We identified 7,852 patients from SEER-Medicare diagnosed with stage IB-IIIA NSCLC from 2000-2013 who underwent lobectomy but did not receive adjuvant chemotherapy. Survival time was calculated from lobectomy date to date of death from NSCLC or other causes. Surviving patients were censored at the time of last known contact. Cause of death was determined from SEER data. The Fine-Gray competing risk model was used to estimate sub-hazards of death from NSCLC vs. other causes; cumulative incidence functions were generated and used to calculate cause-specific survival curves and monthly death rates. The monthly death rates were then incorporated into COLT-M as transition probabilities for each cycle. With this data, we also calculated 5-year overall survival rates, an outcome that is reported commonly in cancer clinical trials. We then modeled until age 100 or death and estimated 10-year overall survival rates for patients with comorbidities who underwent lobectomy followed by adjuvant chemotherapy or observation.

### Treatment-related Complications

COLT-M incorporated the impact of chemotherapy-related adverse events on survival and quality of life. As an increase in non-NSCLC deaths was seen in the first 6 months of follow-up in a pooled analysis of trials evaluating adjuvant chemotherapy (likely as a consequence of complications of treatment), a HR of 2.41 (95% CI: 1.64-3.55) was used in our model to increase the non-NSCLC mortality rate in the first 6 months after adjuvant chemotherapy, after which non-NSCLC mortality returned to background levels.(14) We used pooled data from SEER-Medicare and Kaiser Permanente Southern California (KPSC) to estimate the expected incidence of chemotherapy-associated toxicity conditional on age, sex, and comorbidities, and in the model, non-fatal chemotherapy toxicity was incorporated as a disutility. KPSC includes detailed clinical data including NSCLC characteristics, outcomes, treatment, complications, and comorbidity data that are linked to KPSC’s electronic medical record system. Presence of COPD, CAD, and CHF were ascertained through diagnostic code data from inpatient and outpatient claims files from the 1-year time span prior to being diagnosed with NSCLC.(27, 28) CHF cases were classified as severe if there were hospitalizations for this condition in the year leading up to being diagnosed with NSCLC and were otherwise considered mild/moderate cases. We selected patients with stage IB-IIIA NSCLC who were treated with lobectomy and platinum-based adjuvant chemotherapy (diagnosed 2000-2015 from SEER Medicare and 2005-2015 from KPSC), which yielded 2,301 individuals. We then used diagnostic code data to identify patients who experienced chemotherapy-related complications requiring hospitalization including serious infection, neutropenia, nausea and vomiting, anemia, thrombocytopenia, neuropathy, and renal failure. We fitted logistic regression models to estimate the adjusted probability of developing these complications conditional on age, stage, and comorbidities (COPD, CAD, and CHF) for NSCLC patients receiving adjuvant chemotherapy. The probability of complications in the observation arm were adjusted down applying the relative risk of complications with vs. without adjuvant chemotherapy from the ANITA clinical trial (29).

### Quality-Adjusted Life Expectancy

COLT-M also accounted for quality-adjusted life years (QALYs), an outcome which considers both quality and quantity of life.(30) We derived the negative impact (i.e., disutilities) of treatment-related complications from the literature (31, 32). Quality-of-life parameters for patients with stage IB-IIIA NSCLC and comorbidities were generated using the SEER registry linked to the Medicare Health Outcomes Survey (SEER-MHOS) for a different study (unpublished). MHOS includes self-reported comorbidity data (COPD, CAD, and CHF status). Using a published algorithm, data from MHOS quality-of-life instruments were converted to a summary utility score and utility values were estimated with linear regression analysis according to comorbidity type (33, 34), with estimated utility values -0.018 for CAD, -0.021 for COPD and -0.069 for CHF (Table 1).

### Comparative Effectiveness Analyses: Base Case and Sensitivity Analyses

Gains in QALYs were used to compare patients with comorbidities who received adjuvant chemotherapy vs. observation. Adjuvant chemotherapy was considered the optimal treatment strategy when it was associated with least a 0.25 (3 month) QALY increase. Otherwise, observation was considered the favored strategy. This threshold was selected based on the American Society of Clinical Oncology benchmarks for clinical significance for NSCLC (35).

For the base case analyses, we compared QALY gains and 5-year survival rates of adjuvant chemotherapy vs. observation alone. In these analyses, we used COLT-M to calculate survival and QALYs in patients with stage IB-IIIA NSCLC and comorbidities according to age category (66-69, 70-74, 75-79, and 80-84 years), comorbidity type (COPD, CAD, and CHF), and NSCLC stage (IB, IIA, IIB, and IIIA). We conducted analyses for lone comorbidities (COPD, CAD, and CHF) and combinations of comorbidities (CAD/CHF, CAD/COPD, CHF/COPD, CAD/COPD). We conducted one-way sensitivity analyses, evaluating model output when varying treatment effectiveness and toxicity (from RCTs) parameters over their 95% CIs.

## Results

### Estimates of Lung Cancer Parameters

SEER-Medicare analyses found that each increasing age category was associated with greater risk of non-NSCLC cancer death and female sex was associated with a decreased risk of death from non-NSCLC causes (Table 2; HR: 0.78; 95% CI: 0.70-0.87). The presence of comorbidities was also associated with higher non-NSCLC death risk (COPD HR: 1.18; 95% CI: 1.06-1.31, CAD HR: 1.25; 95% CI: 1.11-1.41, CHF mild/moderate HR: 1.49; 95% CI: 1.21-1.83 and CHF severe HR: 1.88; 95% CI: 1.48-2.37, respectively).

**Table 2.** Adjusted risk factors for death from non-lung cancer causes for patients with stage IB NSCLC treated with lobectomy and no adjuvant chemotherapy (n=7,852)

|  |  |
| --- | --- |
| **Covariates** | **Hazards Ratio**<br>**(95% Confidence Interval)** |
| **Age (Ref: 66-69 years)** |  |
| 70-74 years | 1.17 (1.01-1.37) |
| 75-79 years | 1.33 (1.14-1.55) |
| 80-84 years | 1.62 (1.38-1.92) |
| **Female** | 0.78 (0.70-0.87) |
| **Chronic Obstructive Pulmonary Disease** | 1.18 (1.06-1.31) |
| **Coronary Artery Disease** | 1.25 (1.11-1.41) |
| **Congestive Heart Failure (Ref: No Congestive Heart Failure)** |  |
| Mild/Moderate | 1.49 (1.21-1.83) |
| Severe | 1.88 (1.48-2.37) |

Adjusted logistic regression models of major complications of patients who underwent lobectomy and received adjuvant chemotherapy (Table 3) demonstrated that female sex was significantly associated with nausea and vomiting (HR: 2.33; 95% CI: 1.48-3.71). COPD was a predictor for infection (HR: 1.80; 95% CI: 1.32-2.45), anemia (HR: 1.41; 95% CI: 1.12-1.79), and thrombocytopenia (HR: 2.39, 95% CI: 1.45-3.88). CHF (all severity levels) was associated with an increased risk of nausea and vomiting (HR: 5.58, 95% CI: 2.49-12.49) and thrombocytopenia (HR: 3.46, 95% CI: 1.36-8.78). CHF was also associated with anemia (mild/moderate CHF HR: 2.08; 95% CI: 1.1-3.96; severe CHF: HR: 2.81; 95% CI: 1.5-5.2). CAD was not significantly associated with major complications of adjuvant chemotherapy.

**Table 3.** Adjusted risk factors of major adjuvant chemotherapy complications in lobectomy patients who underwent adjuvant chemotherapy (n=2,301)

| Model | Variables<br>Odds Ratio (95% Confidence Interval) |  |  |  |  |  |
| --- | --- | --- | --- | --- | --- | --- |
|  | Age | Female | COPD | CAD | Mild/Moderate CHF | Severe CHF |
| Serious Infection | NS | NS | 1.80 (1.32-2.45) | NS | NS | NS |
| Neutropenia | NS | NS | NS | NS | NS | NS |
| Nausea and Vomiting | NS | 2.33 (1.48-3.71) | NS | NS | 5.58 (2.49-12.49) |  |
| Anemia | NS | NS | 1.41 (1.12-1.79) | NS | 2.08 (1.1-3.96) | 2.81 (1.5-5.2) |
| Thrombocytopenia | NS | NS | 2.39 (1.46-3.88) | NS | 3.46 (1.36-8.78) |  |
| Neuropathy | NS | NS | NS | NS | NS | NS |
| Renal Failure | NS | NS | NS | NS | NS | NS |
| NS = Not Significant; COPD = Chronic obstructive pulmonary disease; CAD = Coronary artery disease; CHF = Congestive heart failure |  |  |  |  |  |  |

### Assessment of Adjuvant Chemotherapy vs. Observation in Simulated Trials

Using COLT-M, we performed in-silico clinical trials that compared QALYs for adjuvant chemotherapy vs. observation in patients with stage IB-IIIA NSCLC who underwent lobectomy (Figure 1 and S1 Table). We also calculated 5-year survival rates (S2 Table).

**Figure 1.**
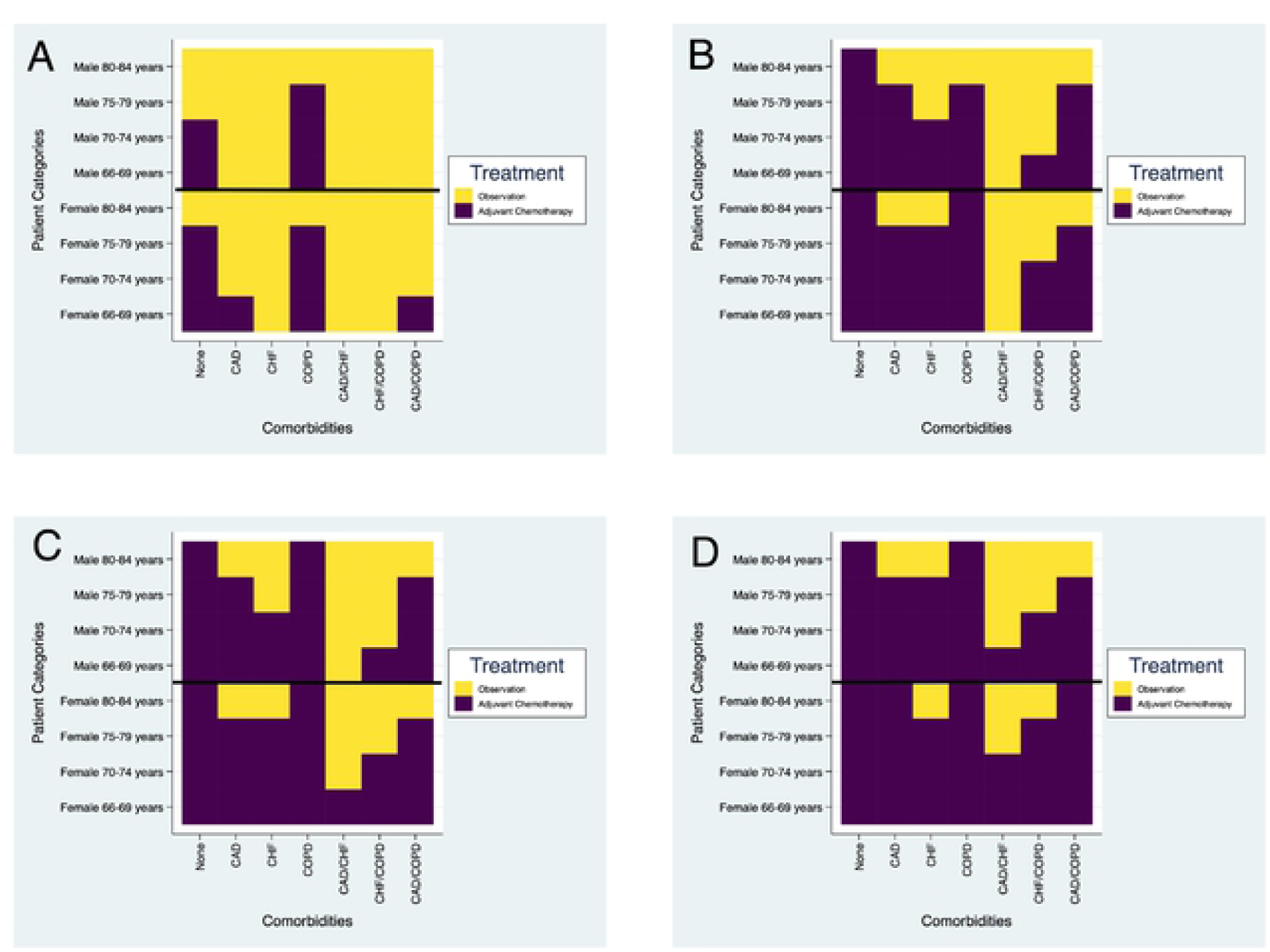
Estimated treatment strategy associated with greatest quality-adjusted life-year gains (QALY) by patient age, sex and comorbidity groups for non-small cell lung cancer (A) stage IB; (B) stage IIA; (C) stage IIB; (D) stage III.

In simulated patients with stage IB NSCLC and comorbidities (Figure 1A), adjuvant chemotherapy was shown to be the optimal predicted therapy (at least a 0.25 (3 month) QALY increase) compared to observation in men 66-79 years old with COPD only (0.25-0.4 QALY gains and led to 2-4% increase in 5-year overall survival). Estimated treatment benefits differed for women; those who were 66-69 years old with CAD or 66-79 years old with COPD derived greatest benefits from adjuvant chemotherapy, with absolute increases ranging from 0.25-0.4 QALYs and 5-year survival increasing by 1-2%. Observation was considered the optimal strategy for all patients >80 years old and all patients with CHF, CAD/CHF, CAD/COPD (except among women aged 66-69), and CHF/COPD.

For stage II NSCLC (Figures 1B and 1C), adjuvant chemotherapy was projected to be the optimal treatment option (>0.25 QALY gains) for more comorbidity groups than for stage IB. In both men and women with single comorbidities (CAD, CHF, and COPD), these included all patients except those in the oldest age categories (0.3-0.6 QALY gains, 2-5% increase in 5-year overall survival). Adjuvant chemotherapy was predicted to be the optimal treatment in fewer patients with CHF/COPD and CAD/COPD (0.3-0.6 QALY gains, 3-4% increase in 5-year overall survival) and observation was the preferred treatment in men and women with CAD/CHF.

For simulated patients with stage IIIA disease (Figure 1D), adjuvant chemotherapy was considered the optimal treatment in the majority of patient groups. The greatest QALY gains were seen with adjuvant chemotherapy in all age categories in patients with comorbid COPD alone (range: 0.25-0.6 QALY gains and increases in 5-year overall survival ranging 4-6%). Most patients in the single comorbidity groups maximally benefited from adjuvant chemotherapy except the oldest groups of men (80-84 years) with CAD or CHF; the oldest group of women (80-84 years) differed slightly, with chemotherapy lacking benefits only for CHF patients. All multimorbid women younger than 75 years had clinically significant benefits with adjuvant chemotherapy (QALY gains ranging 0.3-0.5, 3-4% increases in 5-year overall survival), while observation was associated with non-inferiority in some scenarios (e.g., women aged 75-79 years with CAD/CHF and women 80-84 years old with CHF/COPD). Men aged 66-69 years with CAD/CHF, 66-74 years with CHF/COPD and aged 66-79 years with CAD/COPD were projected to have clinically significant benefits with adjuvant chemotherapy (QALY gains ranging 0.3-0.4, 3-4% increases in 5-year overall survival rates).

Patients without comorbidities were also assessed as validation to show similar results to existing RCTs. Adjuvant chemotherapy was considered the optimal therapy in all patient groups except older patients with Stage I NSCLC (men 75-84 years old and women 80-84 years old).

One-way sensitivity analyses evaluated the robustness of model outputs to changes in key parameters (S3 and S4 Tables).

## Discussion

We conducted simulated clinical trials using the COLT-M, an extension of an established lung cancer microsimulation model, to assess the indications for adjuvant platinum-based chemotherapy in stage IB-IIIA NSCLC patients with comorbidities. Our results showed that the optimal treatment strategy (adjuvant chemotherapy vs. observation) differed by age, sex, and comorbidities. While some patients benefited from adjuvant therapy, some older patients, particularly with earlier stages of disease, had better outcomes with observation after undergoing lobectomy. This study shows that while comorbidities should not prevent the use of adjuvant chemotherapy in many patients, a less aggressive treatment strategy may be the preferred approach for a select group of patients who are older or have major cardiac comorbidities.

Meta-analyses of RCTs and retrospective observational studies have shown that adjuvant chemotherapy for early-stage NSCLC after lobectomy leads to improvements in overall survival, with absolute increases ranging from 4 to 9% (13, 14, 36, 37). These studies have informed clinical guidelines recommending the use of adjuvant platinum-based therapy for locoregional NSCLC (11). While a large proportion of patients with NSCLC have comorbid COPD, CAD, and CHF, these comorbidities are common exclusion criteria from these RCTs and there is limited guidance regarding how to optimally treat these patients (6-10, 15-17). Patients with serious comorbidities warrant special consideration when assessing the harms and benefits of adjuvant chemotherapy given the potential substantial toxicities of therapy, differences in baseline quality of life, and worse overall survival in this patient population (18-22). Patients with comorbidities are also less likely to receive aggressive cancer treatment, which highlights the importance of studying the indications for more intensive treatment modalities in these patients (6).

Simulation modeling is a way of leveraging existing data to assess the risks and benefits of adjuvant chemotherapy and can provide guidance for a wide variety of patient profiles. In our study, simulated patients in COLT-M received treatment with either adjuvant chemotherapy or observation. Our simulated clinical trials considered both quantity and quality of life and compared QALYs between the treatment groups according to patients’ NSCLC stage, sex, age category, and comorbidity profile. In general, patients were more likely to have a greater benefit from adjuvant therapy if NSCLC stage was more advanced. This is consistent with adjuvant therapy clinical trial data and previous observational studies (14, 36). Our study also found that patients >80 years old in many comorbidity categories were less likely to benefit from adjuvant chemotherapy, which is consistent with a previous SEER-Medicare observational study that demonstrated that adjuvant chemotherapy did not have a survival benefit for adults 80 years and older for stage II-IIIA NSCLC (38).

We also found that many patients with COPD benefit from receiving adjuvant chemotherapy. Lung cancer patients with COPD have worse outcomes and are less likely to receive adjuvant therapy (39, 40). Additionally, lung cancer may be more aggressive in patients with COPD (41-43). The results of our study confirm that though patients with COPD are more likely to have major complications from adjuvant chemotherapy and worse lung cancer outcomes, many of these patients have significant gains in QALYs with adjuvant treatment compared to observation alone.

Cardiovascular comorbidities were most frequently associated with lack of clinically significant superiority for adjuvant chemotherapy, particularly in older patients and earlier stage tumors. Our findings were likely driven by substantial increases in non-lung cancer death with these conditions (which attenuates the benefits of chemotherapy), as well as higher treatment toxicity and a negative influence on quality of life. While there is currently very limited guidance on use of adjuvant chemotherapy, patients with lung cancer and comorbid cardiovascular diseases are known to have worse overall survival largely due to competing risks of death (44, 45). Our study’s results support that cardiovascular conditions are an important consideration in weighing the risks and benefits of adjuvant chemotherapy and should play a role in treatment decision-making.

Our analyses showed that female sex was associated with improved survival, worse chemotherapy toxicity, and worse quality of life, findings which are consistent with the existing literature (46-48). In our simulated clinical trials, women with advanced age and more comorbidities benefited more from adjuvant chemotherapy than men in the same groups. Our findings provide additional support that women may benefit from more aggressive NSCLC treatment than men and that the decision to use adjuvant chemotherapy in men, a group with worse lung cancer outcomes who are less likely to benefit from this treatment, should carefully consider age and comorbidity status.

A strength of our study is that we used a robust modeling approach to assess the indications for adjuvant chemotherapy in older patients with locoregional NSCLC and comorbidities. This simulation modeling approach is well-established in evaluating lung cancer screening and treatment and has been used to evaluate treatment benefits of surgery in patients with comorbidities (27, 49-51). Our model leverages data from large, diverse clinical datasets and existing clinical trial and observational data (23). Simulation modeling also has the advantage of assessing the benefit of treatments for a wide variety of patient profiles, allowing clinical guidance for very specific clinical scenarios. However, our study also has limitations that should be discussed. First, we ascertained the presence of comorbidities using diagnostic codes, which may not accurately capture disease and has limitations in evaluating disease severity. However, diagnostic codes for CAD have been shown to very specific in validation studies and we used validated algorithms to enhance the accuracy of diagnosis codes (27, 28, 52). We only evaluated patients with lobectomy, the standard treatment for stage IB-IIIA NSCLC; thus our results do not apply to patients who underwent limited resection that are at increased risk of recurrence (53, 54). Additionally, we did not assess the cost-effectiveness of adjuvant chemotherapy in patients with comorbidities, which may differ from that estimated for healthier populations with locoregional NSCLC (55, 56). We did not assess adjuvant treatment with targeted therapy, which is now indicated for patients with early-stage lung cancer with targetable mutations (11, 57, 58). Similar to RCTs of adjuvant platinum-based therapy, clinical trials evaluating adjuvant targeted therapies excluded patients with severe comorbidities (57, 58). As the uptake increases in real-world populations, we will be able to assess the benefits of targeted adjuvant therapy in patients with comorbidities in the future using the simulation model framework developed for this study.

In summary, we used a simulation model to evaluate QALY increases from adjuvant chemotherapy for patients with stage IB-IIIA resected NSCLC and comorbidities. We found that many older patients with multiple comorbidities can benefit from adjuvant therapy; however, older individuals with cardiac comorbidities may be better managed with observation. Our simulation model results help to extend existing clinical trial data to unstudied, high-risk populations for whom limited primary data exist and may reduce clinical uncertainty regarding the use of adjuvant chemotherapy. Our study findings can be used to guide future research on treatment in patients with early-stage NSCLC and comorbidities, such as studies on adjuvant targeted therapies and immunotherapy.

## Data Availability

SEER-Medicare data is managed by the Centers for Medicare and Medicaid Services. Although personal identifiers for all patient and medical care providers have been removed from the SEER-Medicare data, data cannot be shared publicly as there remains the remote risk of re-identification (given the large amount of data available). Data can be accessed, subject to approval and data use agreement, from the Healthcare Delivery Research Program at the National Cancer Institute (http://appliedresearch.cancer.gov/seermedicare/obtain/requests.html).The data used for this study from Kaiser Permanente Southern California (KPSC) contain protected health information (PHI) and access is protected by the Kaiser Permanente Southern California (KPSC) Institutional Review Board (IRB) and owned by KPSC. For more information about data access and the criteria for access to confidential data, please contact the study author and the KPSC IRB.

10.6084/m9.figshare.19086659

## Acknowledgements

The authors acknowledge the efforts of the National Cancer Institute; the Office of Research, Development and Information, CMS; Information Management Services (IMS), Inc.; and the Surveillance, Epidemiology, and End Results (SEER) Program tumor registries in the creation of the SEER-Medicare database. The interpretation and reporting of these data are the sole responsibility of the authors.

## Supporting Information

**S1 Table**. Estimated quality-adjusted life expectancy according to patient group, treatment and comorbidity.

**S2 Table**. Estimated 5-year survival rates according to patient group, treatment and comorbidity

**S3 Table**. Percent of scenarios changing conclusions in sensitivity analyses: Varying lung cancer survival benefit over 95% confidence interval of hazard ratio

**S4 Table**. Percent of scenarios changing conclusions in sensitivity analyses: Varying post-chemotherapy mortality penalty

